# Deep Learning based Intraretinal Layer Segmentation using Cascaded Compressed U-Net

**DOI:** 10.1101/2021.11.19.21266592

**Authors:** Sunil K. Yadav, Rahele Kafieh, Hanna G. Zimmermann, Josef Kauer-Bonin, Kouros Nouri-Mahdavi, Vahid Mohammadzadeh, Lynn Shi, Ella M. Kadas, Friedemann Paul, Seyedamirhosein Motamedi, Alexander U. Brandt

## Abstract

Intraretinal layer segmentation on macular optical coherence tomography (OCT) images generates non invasive biomarkers querying neuronal structures with near cellular resolution. While first deep learning methods have delivered promising results with high computing power demands, a reliable, power efficient and reproducible intraretinal layer segmentation is still an unmet need. We propose a cascaded two-stage network for intraretinal layer segmentation, with both networks being compressed versions of U-Net (CCU-INSEG). The first network is responsible for retinal tissue segmentation from OCT B-scans. The second network segments 8 intraretinal layers with high fidelity. By compressing U-Net, we achieve 392- and 26-time reductions in model size and parameters in the first and second network, respectively. Still, our method delivers almost similar accuracy compared to U-Net without additional constraints of computation and memory resources. At the post-processing stage, we introduce Laplacian-based outlier detection with layer surface hole filling by adaptive non-linear interpolation. We trained our method using 17,458 B-scans from patients with autoimmune optic neuropathies, i.e. multiple sclerosis, and healthy controls. Voxel-wise comparison against manual segmentation produces a mean absolute error of 2.3*μ*m, which is 2.5x better than the device’s own segmentation. Voxel-wise comparison against external multicenter data leads to a mean absolute error of 2.6*μ*m for glaucoma data using the same gold standard segmentation approach, and 3.7*μ*m mean absolute error compared against an externally segmented reference data set. In 20 macular volume scans from patients with severe disease, 3.5% of B-scan segmentation results were rejected by an experienced grader, whereas this was the case in 41.4% of B-scans segmented with a graph-based reference method.

## 1 Introduction

Optical coherence tomography (OCT) is a noninvasive in-vivo imaging modality, which is able to acquire 3D volume scans of the retina with micrometer resolution (Huang et al. (1991)). The retina is part of the central nervous system, and OCT-derived intraretinal layer thickness or volume measurements, which are typically measured within the macular region, are promising non invasive biomarkers querying neuronal structures. For instance, the combined macular ganglion cell and inner plexiform layer (GCIPL) shows thinning in eyes affected by optic neuritis (ON), multiple sclerosis (MS) (Pawlitzki et al. (2020)), neuromyelitis optica spectrum disorders (NMOSD) (Filippatou et al. (2020)) and Alzheimer’s disease (Cabrera et al. (2019)). GCIPL was also shown to be associated with future disease activity in patients with clinically isolated syndrome (CIS) suggestive of MS (Zimmermann et al. (2018)) and of cognitive decline in Parkinson’s disease (Murueta-Goyena et al. (2021)). The macular inner nuclear layer (INL) may be affected by inflammatory activity in MS and related disorders (Balk et al. (2019)) or Müller cell damage in NMOSD (Oertel et al. (2017); Motamedi et al. (2020)). In glaucoma, thinning of inner retinal layers such as GCIPL and retinal nerve fiber layer (RNFL) often appears before visual function loss and may be useful as a biomarker for early diagnosis (Mohammadzadeh et al. (2020)). Notably, these applications rely on changes in layer thickness, while the overall configuration of the retina is mostly unaltered, which separates their segmentation needs from disorders with clear microscopic retinal pathologies such as age-related macular degeneration (AMD) or diabetic retinopathy (DR). Consequently, intraretinal layer segmentation is not a trivial task as layer changes can be small, while texture gradients can be weak and influenced by noise (Oberwahrenbrock et al. (2016)). While many OCT manufacturers provide device-integrated segmentation of some intraretinal layers, results are often noisy and regularly require manual correction, which in itself is challenging for clinical use and difficult to handle in research settings when multiple graders review macular OCT images (Oberwahrenbrock et al. (2016, 2018)). A reliable and robust algorithm for intraretinal layer segmentation that requires no or minimal manual correction is an unmet need to foster reliable and reproducible research as well as clinical application (Cruz-Herranz et al. (2016)).

In this paper, we propose a cascade of compressed convolutional neural networks with a U-Net like architecture for fully-automated intraretinal layer segmentation (CCU-INSEG). We compress the U-Net models to reduce required resources without losing a significant amount of accuracy in the segmentation process. The cascaded architecture helps the algorithm increase robustness with regard to different type of OCT images without adding too many parameters in the overall network. We validate the method using internal and external data on a voxel, measurement parameter and scan level.

### 1.1 State of the Art

Intraretinal layer segmentation of OCT-derived macular volume scans can be divided into two categories: Mathematical modelling-based methods and machine learning (ML)-based methods, with the latter recently extending towards deep learning.

Mathematical modelling-based methods follow a set of rules based on well defined deterministic mathematical and image analysis concepts to segment intraretinal layer boundaries. These methods are not within the scope of this paper and we would like to refer readers to a recent state-of-the-art review summarizing these approaches (DeBuc (2011)).

Data-driven or ML-based methods learn features from image data sets automatically and have produced reliable and accurate measurements in several retinal imaging applications (Ting et al. (2019)). In the first paper proposing deep learning derived methods for intraretinal layer segmentation, Fang et al. (2017) used a Cifar-style convolutional neural network (CNN) to create class labels and probability maps, followed by a graph theory and dynamic programming step to derive final layer boundaries in OCT images from patients with age-related macular degeneration (AMD). Shortly thereafter, Roy et al. (2017) proposed an end-to-end fully convolutional framework to perform intraretinal segmentation, which was also able to delineate fluid collection in diabetic retinopathy. The authors avoided all deterministic pre- and postprocessing and performed the entire segmentation procedure using a fully convolutional network inspired by DeconvNet (Shelhamer et al. (2017)) and U-Net (Ronneberger et al. (2015)) with an encoder-decoder architecture, skip connections and unpooling layers. Kugelman et al. (2018) used recurrent neural networks (RNN) instead CNNs for 2D image segmentation by formulating image patches as a sequential problem, as previously suggested by ReNet. RNNs have been mainly developed for tasks involving sequential data; in image analysis examples include time series or image stacks. Unlike CNNs, RNNs use interim output as input in the next iteration of computations, thereby utilizing a memory network in a specific classification task on sequential data (Chen et al. (2015)).

Several recent publications make use of the U-Net design, which was introduced to target problems in medical image analysis (Ronneberger et al. (2015)). U-Net’s symmetrical architecture consists of a contracting path to capture context and an expanding path that enables precise localization, linked by skip connections. Mishra et al. (2020) proposed a method similar to Fang et al. but relying on U-Net for generation of probability maps and shortest path segmentation for final boundary detection. He et al. (2019) proposed an intraretinal segmentation method using 2 cascading U-Nets, the first one preforming the actual intraretinal segmentation task, the second one then reconstructing 3D surfaces. Combining segmentation with surface reconstruction using CNN was initially suggested by Shah et al. (2018) and has the benefit that incorrect segmentation results in 2D panes are corrected when reconstructing a continuous surface. This approach has been recently combined into a single unified framework by the same group (He et al. (2021)). Pekala et al. (2018) proposed a method based on DenseNet, which makes use of more extensive skip connections but is otherwise similar in design to U-Net for boundary pixel segmentation, followed by a Gaussian process regression for surface extraction. Another interesting extension of the U-Net concept for retinal segmentation was recently proposed by Liu et al. (2020), who used a nested U-Net shape for multi-scale input and output; this approach performed favorably based on accuracy in comparison to traditional U-Net architectures. Instead of U-Net, Li et al. (2020) proposed a method utilizing a combination of Xception65 network and an atrous spatial pyramid pooling module for retinal boundary detection.

Segmentation accuracy of these previously proposed deep learning-based methods is favourably, with more recent and complex approaches performing better than early methods utilizing simpler network architectures. Still, apparent differences in segmentation performance may rather result from different training data set sizes, additional retinal features like AMD or DR complicating segmentation, and from differing validation strategies. Hamwood et al. (2018) showed that network design choices like patch size may have a strong influence over network performance in retinal segmentation tasks. In contrast, in real world medical applications, selection of training data, meaningful pre- and postprocessing based on domain knowledge, and computational costs are highly relevant. Especially the latter heavily favors simpler network architectures, because complex architectures with multiple parameters are highly resource intensive, which is prohibitive for environments with limited resources. Methods implementing lean networks and specific post processing for segmentation errors, e.g. by reconstructing surfaces, can achieve high segmentation efficiency while being performant (He et al. (2021)).

One way performance optimization may be achieved is by compressing network architectures, for example by architecture pruning, weight quantization or knowledge distillation (Mangalam and Salzamann (2018)). The latter was recently used by Borkovkina et al. (2020) to propose a retinal segmentation method, which utilizes a performance-optimized smaller network that was trained on a complex original segmentation network as teacher network.

As our main contribution, we here show that compression of a U-Net style network based on architecture pruning leads to substantial performance improvements while not suffering in accuracy. We further show that appropriate pre- and post-processing is beneficial to reduce errors and achieve high segmentation accuracy, introducing an adaptive hole filling process. We further introduce a framework for method validation based on the clinically more relevant computed thickness and volume parameters instead of voxel-wise comparisons alone. Of note, our method was also trained on more diverse data than previous approaches, improving its readiness for immediate application rather than just being a methodological proposal. Together, these contributions highlight important pathways from research-driven implementations towards application.

## 2 Material and Methods

The proposed method starts with pre-processing an input volume scan to remove undesired components from each B-scan (Figure 1). It then performs intraretinal segmentation using a cascaded network, where each stage is a compressed U-Net like network. Finally, during post-processing we derive continuous surfaces and introduce anatomical constraints. In the following, we explain each component of the pipeline in detail.

**Figure 1:**
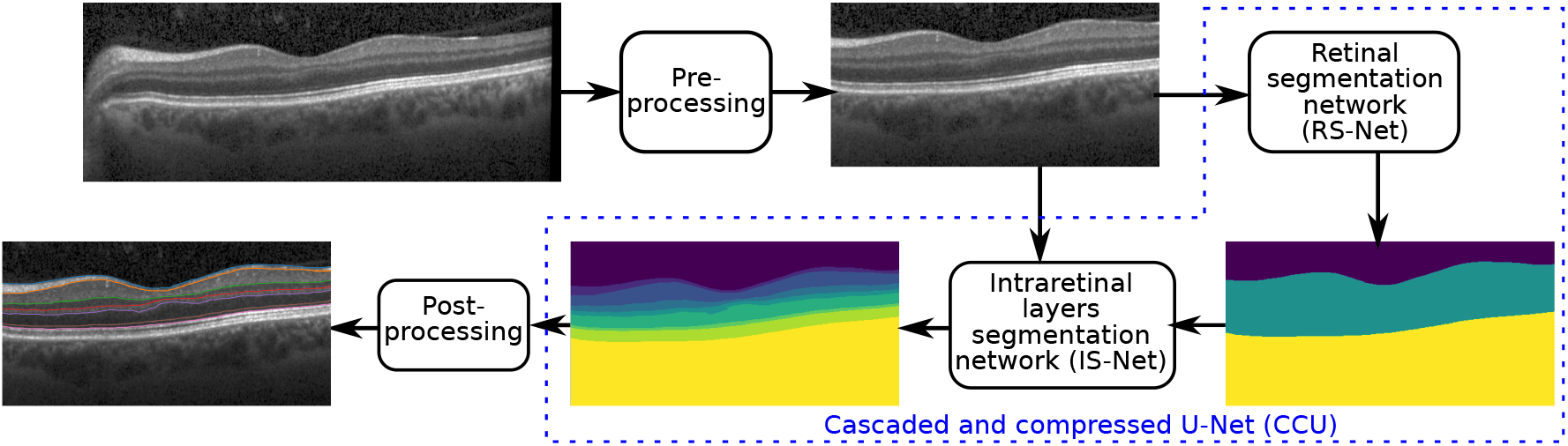
The pipeline of the proposed method consists of four steps: pre-processing, retinal region segmentation, layer segmentation, and post-processing.

### 2.1 Segmentation Method

#### 2.1.1 Pre-processing

Let us consider an input OCT volume scan sampled from the macula consisting of several B-scans. During data acquisition, artefacts are inevitable due to various internal and external factors. These artefacts may include noise, which is responsible for low signal-to-noise (SNR) ratio, cut scans (upper and lower cuts), and improper focus (Tewarie et al. (2012)). Additionally, parts of the optic nerve head (ONH) frequently appear in macular scans as shown in Figure 1 (the input image). These artefacts are not desirable and affect the performance of segmentation algorithms.

To remove these artefacts from the OCT volumes, we employ a deep learning-based quality check method introduced and described in detail by Kauer et al. (2019). The output of the quality analysis is combined into three key features; centering, signal quality and image completeness. The centering information confirms the focus of scanning, signal quality provides the signal to noise ratio, and image completeness ensures that the input volume is free from cuts. Besides, this method localizes the center of the macula using parabola fitting in the foveal region. Furthermore, based on the computed foveal center, B-scans are cropped from both sides to cover only a central 6×6mm ROI for both training and prediction to avoid undesired ONH region.

#### 2.1.2 Cascaded and Compressed U-Net-style networks

Our segmentation method consists of two cascaded U-Net like architecture as shown in Figure 1. The first network termed RS-Net (retinal segmentation network) is responsible for segmentation of the retina from a full OCT B-scan. In its simplest forms this is the retinal tissue between inner limiting membrane (ILM) and Bruch’s membrane (BM) as boundaries. The trained network takes a pre-processed OCT B-scan as input and produces a three class output: purple region (above ILM), green region (retinal region), and yellow region (below BM) (Figure 1). The output of RS-Net and the original input serve as the input for the second network. The second network, intraretinal layer segmentation network (IS-Net) takes the two channel input and generates a 9-class output providing boundaries for 8 intraretinal layers (Figure 2).

**Figure 2:**
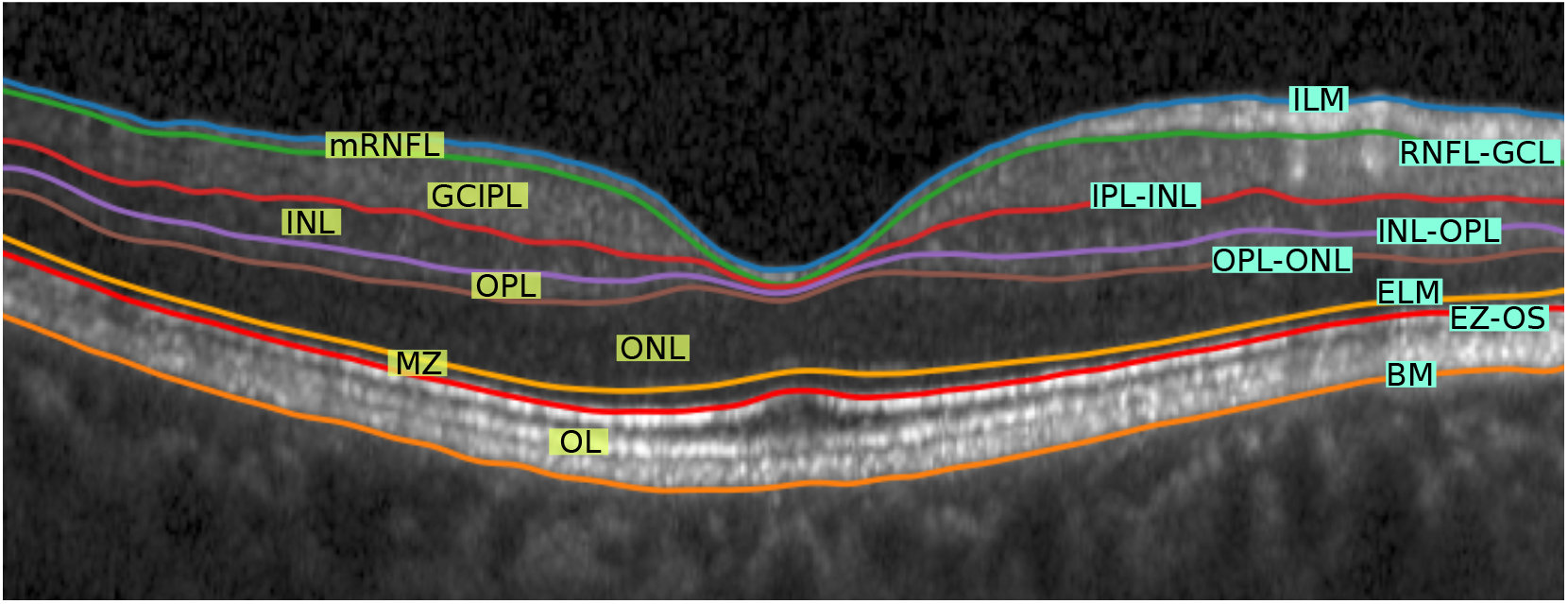
Visual representation of different retinal layers and boundaries after segmentation. Abbreviations: ILM - Inner limiting membrane, mRNFL - Macular retinal nerve fibre layer, GCL - Ganglion cell layer, IPL - Inner plexiform layer, GCIPL - Macular ganglion cell/inner plexiform layer, INL - Inner nuclear layer, OPL - Outer plexiform layer, ONL - Outer nuclear layer, ELM - External limiting membrane, MZ - Myoid zone, EZ - Ellipsoid zone, OS - Outer segment, RPE - Retinal pigment epithelium, OL - Outer layer, and BM- Bruch’s membrane.

The original U-Net is a fully CNN that consists of a contracting path (encoder) and an expansive path (decoder) (Ronneberger et al. (2015)). Paths are connected at multiple places by skip connections, a key aspect of U-net that allows the network to restore spatial information, which were lost during the pooling operation. In general, U-Net is capable of producing high fidelity segmentation results and most of the state-of-the-art segmentation methods are based on U-net architecture and focused on high accuracy (see above). But the deep architecture of U-net requires considerable computational resources, which restricts the deployment of the model on systems with limited processing power. To find a desirable balance between processing needs and accuracy, we studied compressed variations of this network with regard to channel depth and filter size.

Figure 3 shows the architecture of the proposed compressed U-Net. Similar to U-Net, the proposed method has contracting and expansive paths. The contracting or encoder part has four stages and at each consecutive stage, the channel depth gets double and the input size gets half (via max pooling). Additionally, dropout layers are added after the third and the fourth stages to avoid over-fitting. In the decoder part, the inverse of the encoder part is happening except it has two additional blocks: merging and output convolution layers with kernel size of 1 as shown in Figure 3. The terms *h* and *w* represent the size of the pre-processed image. The number of the input channels is represented by *n* and it is clear from Figure 1 that *n* = 1 for RS-Net and *n* = 2 for IS-Net. Optimal architecture parameters values (*h, w, f, c*) are explained in Section 2.3.

**Figure 3:**
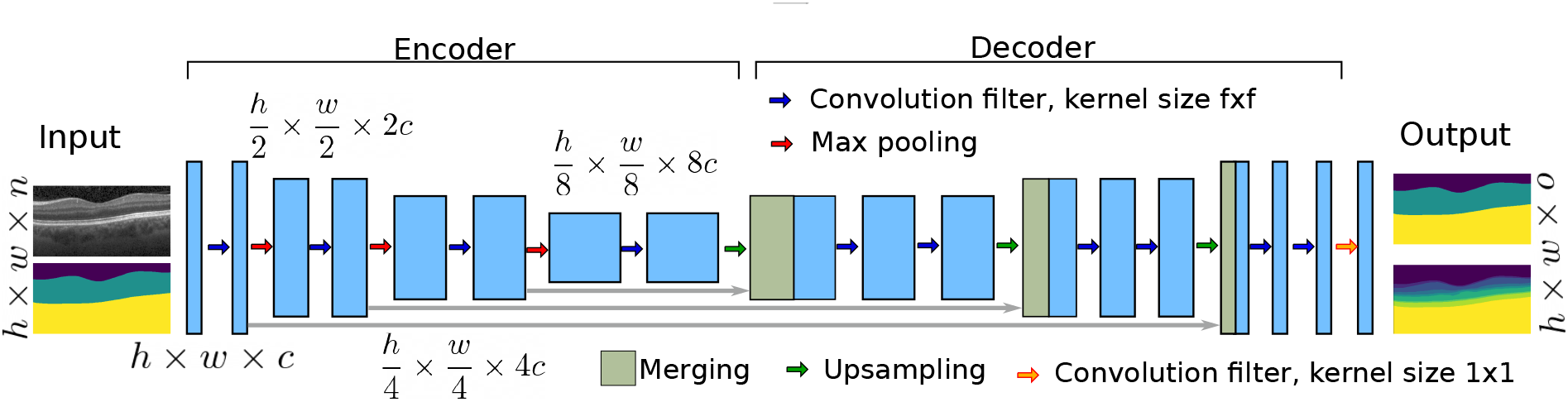
The general architecture of the proposed compressed U-Net. RS-net and IS-Net both have the same architecture with different channel depth *c*, kernel size *f*, number of input channel *n*, and number of the output classes *o*.

**Figure 4:**
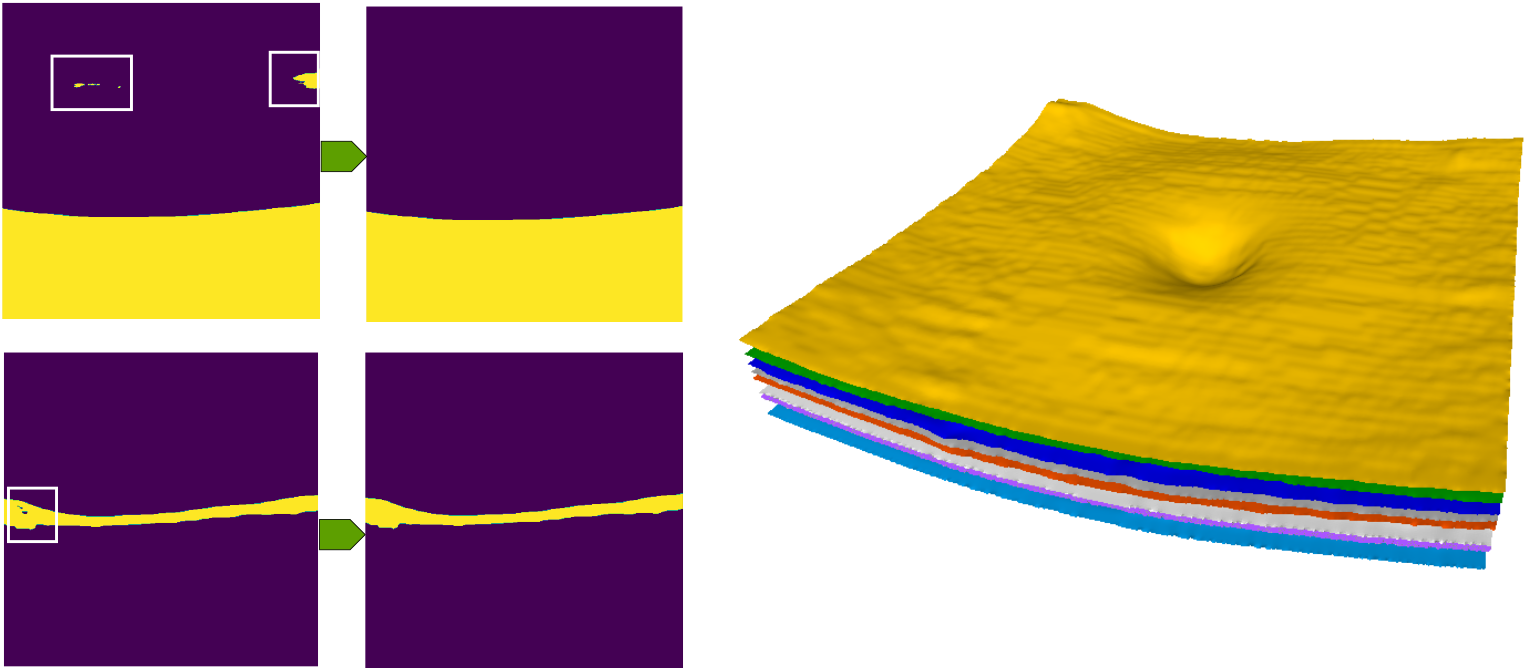
The first column shows small holes, which appear during the classification in both networks. The second column consists of images without holes. The last column shows 3D reconstructed surfaces of different layers.

#### 2.1.3 Loss function

For end-to-end training of the proposed network, we use a combination of categorical cross entropy (CCE) and Tversky as the loss function:

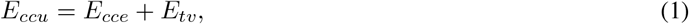

where *E*_*cce*_ represents cross categorical entropy loss function and defined as:

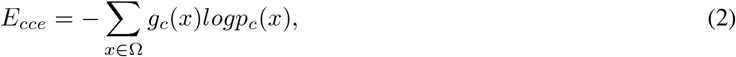

where *x* is the pixel value from the retinal region Ω. The terms *g*_*c*_(*x*) and *p*_*c*_(*x*) show the ground truth and the predicted probability at *x* from the class *c* respectively. The second term of Equation 1 (*E*_*tv*_) represents the Tversky loss function (Salehi et al. (2017)) and addresses the issue of highly imbalanced data as OCT images have thin layers and a wide background:

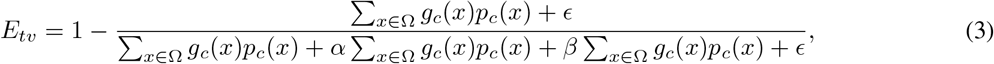

where *g*_*c*_(*x*) = 1 − *g*_*c*_(*x*), *p*_*c*_(*x*) = 1 − *p*_*c*_(*x*), and term *ϵ* is a small constant to avoid division by zero. The second and third terms in the denominator of the above equation are responsible for False Positive (FP) and False Negative (FN) during the pixel classification. Because of ROI imbalance, retinal layer segmentation is sensitive to False Negatives (FN) and Tversky similarity index provides a weight-controlled emphasis on FN and FP by using *α* and *β* as corresponding weighting factor. Salehi et al. (2017) used various statistical analyses and concluded that the network has a balanced output in terms of sensitivity and specificity when *α* = 0.7 and *β* = 0.3 as *α* + *β* = 1. Both networks (RRS-Net and RLS-Net) are trained independently using the above loss function with *α* = 0.7, *β* = 0.3 and *ϵ* = 0.1 respectively.

#### 2.1.4 Post-processing

At the last stage of the pipeline, we apply post-processing on the segmentation outcome to remove artefacts. First, **adaptive hole filling** removes small holes from the background and the foreground of the segmentation outcome. These holes can rarely appear because of the mis-classification of individual or few pixels during the segmentation process. To remove small holes, we use morphological operators with an adaptive threshold. The value of threshold is computed differently for each class of the network’s outcome. RS-Net generates three-class output and for upper and lower classes (above ILM and below BM), the threshold value is *n*_*n*_*/f*, where *n*_*n*_ is the number of non-zero pixels in that class image and *f* is a constant factor with a default value of 2. Additionally, the value of *f* will keep increasing by 1 until *n*_*n*_*/f* < *n*_*z*_, where *n*_*z*_ is the number of zero pixels. The same process is repeated for the upper and lower class of the IS-Net outcome. For the internal layer classes of the networks, the default value of *f* is 10 and it is increased by 1 until *n*_*n*_*/f* < *n*_*z*_.

After holes filling, different layers from the multi-channel outcome of IS-Net are extracted. To detect the boundary in each class image, we search the first non-zero entry in each column. After layer extraction, we remove outliers from the layer surfaces based on Laplacian of the thickness values. Furthermore, we recompute missing values using Piecewise Cubic Hermite Interpolating Polynomial (PCHIP) interpolation. Lastly, an isotropic smoothing with 3 × 3 kernel size is applied to each layer surface to remove small ripples and to generate smoother boundaries.

#### 2.1.5 Implementation

The method was fully implemented in Python. Network training was performed with Python (v3.6) using Keras (v2.3), and TensorFlow (v2.2) libraries on a Linux server with two Intel Xeon E5-2650 CPUs and two NVIDIA GeForce GTX1080 TI GPUs.

### 2.2 Data

To train the proposed method, we selected macular OCT volume scans that were obtained with Spectralis spectral-domain OCT (Heidelberg Engineering, Heidelberg, Germany). All data were selected from the institute’s OCT image database at Charité – Universitätsmedizin Berlin (CUB).

In our previous experience with intraretinal layer segmentation, we noticed that macular OCT volume scans with low, yet acceptable, signal to noise ratio as well as scans from patients with very thin RNFL needed extensive manual segmentation correction. Hence, from a pool of macular scans from healthy controls and patients with CIS, MS, and NMOSD we selected a total of 445 OCT volume scans of 254 eyes, the majority of which with low to average signal to noise ratio, thin RNFL, and microcystic macular pathology, an occasional OCT finding in eyes from patients with various forms of optic neuropathy such as autoimmune optic neuropathies (Kaufhold et al. (2013)). All selected volumes were centered on the fovea covering 25° × 30° with different resolutions (61/25 B-scans, and 768/1024/1536 A-scans per B-scan).

All scans underwent automatic quality control to avoid data with insufficient quality for training and validation purposes (Kauer et al. (2019)). To delineate different OCT layers in these data, the initial segmentation was performed using the SAMIRIX toolbox (Motamedi et al. (2019)), which uses AURA developed by Lang et al. (2013), and then manually corrected by experts using the same toolbox. In the end, we selected 17,458 B-scans for training, 3,081 for validation, and 208 for testing.

#### 2.2.1 Validation data

To validate the data in a multicenter format, additional unseen macular volume scans from our center (CUB) as well as two other centers, University of California, Los Angeles (UCLA) and Johns Hopkins University (JHU) were added.

##### CUB

Twenty-five additional unseen macular 3D OCT scans of 24 eyes were included in this study from our center (CUB). The volumes were from HCs and patients with MS and NMOSD, covering 25° × 30° macular area using 61 B-scans and 768 A-scans per B-scan. All volumes underwent segmentation and manual correction using the SAMIRIX toolbox (Motamedi et al. (2019)). Two scans were rejected by our quality check because of the low signal to noise ratio.

##### UCLA

In order to ensure the quality of the developed method on OCT images from patients with other optic neuropathies, which were not part of the training data, we included a second data set for testing from University of California, Los Angeles (UCLA). The data set consists of 12 OCT volume scans of 12 eyes from glaucoma patients or healthy subjects, each with 61 horizontal B-scans and 768 A-scans per B-scan, covering 25° × 30° centered on the fovea. The OCT volumes in this data set were segmented and then manually corrected using the SAMIRIX toolbox for comparison against the developed method. The volume scans in this data set were cropped to 5.5×5.5 mm square in the pre-processing step to ensure complete exclusion of ONH from macular scans.

##### JHU

To validate our method further, we applied the developed segmentation method on a publicly available OCT dataset from Johns Hopkins University (JHU) (He et al. (2019)). Fifteen volume scans of 15 eyes were selected from this data set, which are a mix of HC and MS data. These scans were segmented and manually corrected by its publisher, so we compared our segmentation results against their manual delineation. The volume scans in the JHU data set cover 20° × 20° of the macula using 49 B-scans and 1024 A-scans per B-scan.

### 2.3 Optimization

To get optimal values of the filter kernel size *f* and the channel depth *c*, we have performed the compression with different values of *f* and *c* as shown in Table 1. The channel depth of the network should be proportional to the complexity of task. From Figure 1, it can be seen that RS-Net produces three channel output (*o* = 3), which is a less complicated task compared to generating 9-class output (IS-net).

**Table 1:**
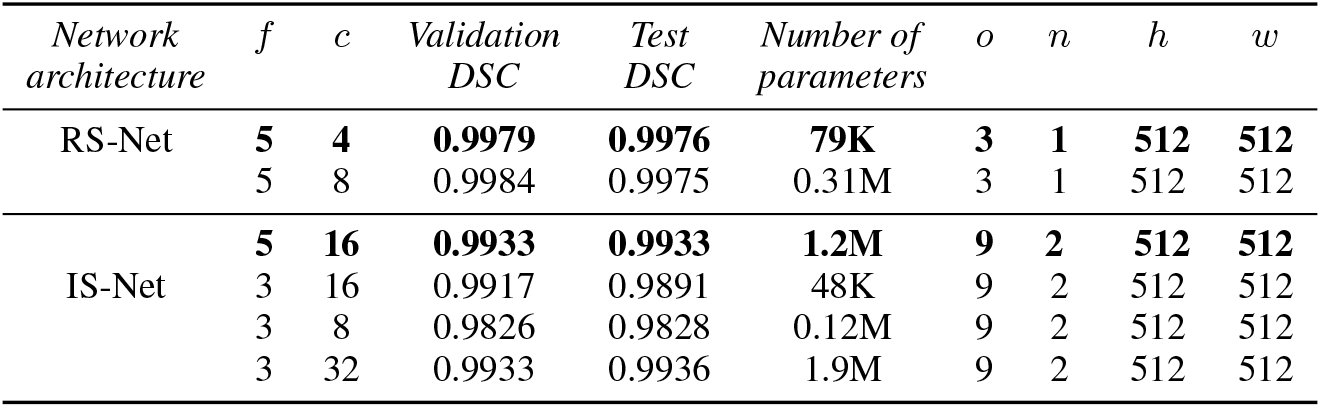
The compression experiments with different architectural parameters for both RS and IS networks.

| Network architecture | $f$ | $c$ | Validation DSC | Test DSC | Number of parameters | $o$ | $n$ | $h$ | $w$ |
| --- | --- | --- | --- | --- | --- | --- | --- | --- | --- |
| RS-Net | **5** | **4** | **0.9979** | **0.9976** | **79K** | **3** | **1** | **512** | **512** |
|  | 5 | 8 | 0.9984 | 0.9975 | 0.31M | 3 | 1 | 512 | 512 |
| IS-Net | **5** | **16** | **0.9933** | **0.9933** | **1.2M** | **9** | **2** | **512** | **512** |
|  | 3 | 16 | 0.9917 | 0.9891 | 48K | 9 | 2 | 512 | 512 |
|  | 3 | 8 | 0.9826 | 0.9828 | 0.12M | 9 | 2 | 512 | 512 |
|  | 3 | 32 | 0.9933 | 0.9936 | 1.9M | 9 | 2 | 512 | 512 |

For RS-Net, we start with two different values of the depth channel (*c* ={4, 8}), which increases up to four times of the input size as shown in Figure 3. RS-Net is responsible for retinal segmentation, and it should not care about tiny details. Therefore, we set kernel size to 5 × 5 (*f* = 5), which helps the network focus on significant features. To check the performance of the RS-Net with different values of *c*, we use a smaller data set (1000 B-scans) for training, validation, and testing along with the loss function, which is explained in Section 2.1.3. As it can be seen from the Table 1, the performance of RS-Net is similar in terms dice similarity coefficient (DSC) for both values of *c*. However, the number of model parameters are almost four times less with *c* = 4 in comparison to *c* = 8. Therefore, *f* = 5 and *c* = 4 are the optimal configuration for RS-Net with respect to model size and performance metrics.

In contrast to RS-Net, IS-Net is responsible for separating 8 layers (9 classes) with the need to detect small details. Different configurations of IS-Net along with the performance metrics are shown in Table 1. The two combinations of channel depth and kernel size (*f, c* ={5, 16} & *f, c* ={3, 32}) produce the best performance metrics. However, the first configuration (*f* = 5, *c* = 16) has a smaller number of model parameters and therefore, is the preferable configuration of IS-Net for our purpose.

In contrast to U-Net, the proposed architecture has fewer stages. U-net consists of 5 stages ranging from 64 to 1024 channel depths and our networks consist of only 4 stages with variable channel depths. Table 2 shows the direct comparison with U-Net and deep residual U-Net (Zhang et al. (2018)) in terms of the number of model parameters, DSC, and the prediction time. ResU-Net performs slightly better compared to the original U-Net and our proposed method. It is worth mentioning that the above comparison is performed at the architectural level. If U-Net will be used in the cascaded setting like Kingma and Ba (2014) then the computational complexity will be high. Although, ResU-Net is used in the cascaded setting so far in the state-of-the-art methods, it has better potential because of the low number of parameters and slightly better performance compared to the original U-Net. However, it still has more parameters than the proposed architecture.

**Table 2:**
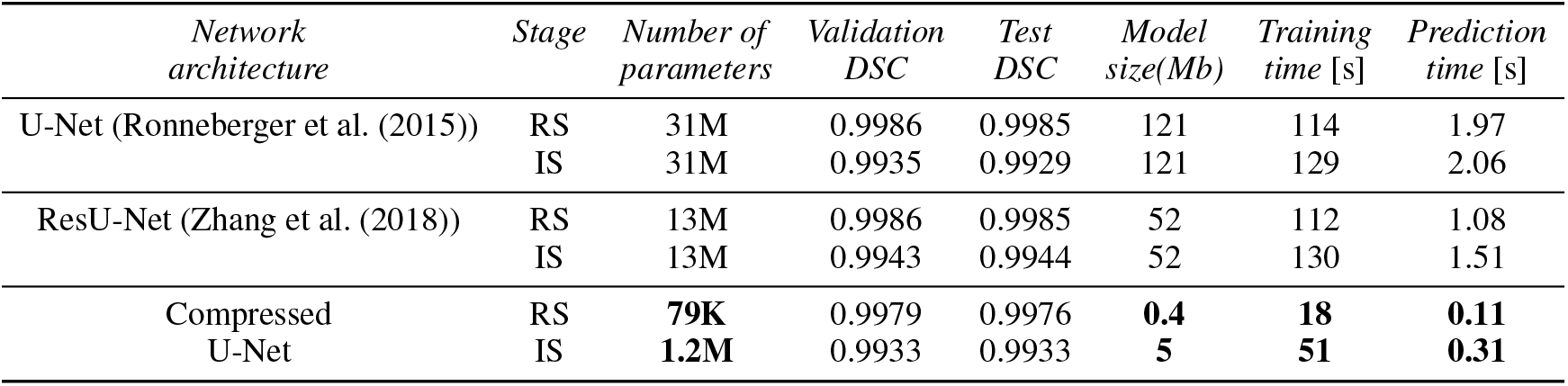
A comparison between the original U-Net and the compressed U-Net (proposed). The prediction time is observed on CPU only for a single B-scan.

As a result, RS-Net is almost 392-times smaller compared to original U-net in terms of parameters and model size. Similarly, IS-Net is almost 26-times smaller compared to U-Net. The comparison of prediction time could be tricky because it depends on several factors including hardware, programming style, libraries, etc. However, it is clear that the smaller network will need smaller computational resources and will be faster compared to U-Net from the running time perspective. Table 2 shows training time for 1 epoch and the mentioned time is taken on a Linux server with two Intel Xeon E5-2650 CPUs and two NVIDIA GeForce GTX1080 TI graphical processing units (GPUs). Furthermore, the prediction time is recorded only on a Core-i7 CPU.

Compared to two-stage cascaded state-of-the-art methods, our CCU network is much smaller regarding parameters and model size with almost similar or better accuracy. For example, Ma et al. (2020) proposed a cascade of two U-Net like architecture, where each network is a combination of original U-Net with additional layers. Therefore, the number of parameters in each network is more than 31M (for the input image size of 512×512), which is much bigger compared to our CCU network. Similarly, He et al. (2019) used a modified U-Net for the first network with a double channel depth compared to our CCU network. The second network of this method consists of two parts: original U-Net and a fully connected final layer. Furthermore, this network is running on patches (8 patches for each B-scan), which also increases the running time complexity.

### 2.4 Statistical Analysis

Intraclass correlation coefficient (ICC) and lower and upper confidence intervals were calculated based on the variance components of a one-way ANOVA, using the ICC package of R. Mean absolute error (MAE), standard deviation, and DSC are computed using Python.

### 2.5 Ethics statement

The study was conducted in accordance with the Declaration of Helsinki in its current applicable form and the applicable European and German laws and was approved by the local ethics committee at Charité – Universitätsmedizin Berlin. All participants gave written informed consent.

## 3 Results

### 3.1 Training Analysis

End-to-end training was performed using 17,458, 3,081, and 208 B-scans for training, validation, and test, respectively. To minimize the loss function (Equation 1), Adam optimizer was used with a fixed learning rate of 0.001. We set the number of iterations to 80 for both networks with early stopping criteria. For early stopping, we monitored the validation accuracy.

As it can be seen from Table 3, the optimal and stable accuracies are reached in 23 and 11 epochs for RS-Net and IS-Net respectively. Additionally, Table 3 consists of different training-testing related metrics of the proposed CCU network.

**Table 3:** Training related metrics. Abbreviations: Train. - Training, Val. - Validation, Acc. - Accuracy

| *Network arch.* | *Train. acc.* | *Train. DSC* | *Train. loss* | *Val. acc.* | *Val. DSC* | *Val. loss* | *Test acc.* | *Test DSCS* | *Test loss* | *Epoch early stop* |
| --- | --- | --- | --- | --- | --- | --- | --- | --- | --- | --- |
| RS | 0.9985 | 0.9992 | 0.0064 | 0.9984 | 0.9992 | 0.0064 | 0.9985 | 0.9991 | 0.0061 | 28 |
| IS | 0.9953 | 0.9974 | 0.0175 | 0.9952 | 0.9974 | 0.0179 | 0.9950 | 0.9973 | 0.0185 | 11 |

### 3.2 Comparison against device-implemented segmentation

We then compared the proposed method and standard device segmentation of Heidelberg Eye Explorer (HeyEx) without further correction. Table 4 shows MAE(SD) between manually corrected gold standard segmentation (15 volume scans of 15 eyes) and the device segmentation (first row). Similarly, the second row shows MAEs(SD) between the proposed method’s outcome and gold standard segmentation. Our method has almost 2.5 times lower MAEs and SDs compared to the device segmentation.

**Table 4:** A comparison between HeyEx segmentation and the proposed method (CCU-INSEG) using MAE (standard deviation) in *μ*m.

| *Method* | *ILM* | *RNFL-GCL* | *IPL-INL* | *INL-OPL* | *OPL-ONL* | *ELM* | *EZ-OS* | *BM* | *Total* |
| --- | --- | --- | --- | --- | --- | --- | --- | --- | --- |
| HeyEx | 3.5(3.2) | 6.3(2.6) | 6.6(2.5) | 7.6(2.5) | 4.8(3.0) | 3.5(2.8) | 5.2(2.9) | 4.0(2.7) | 5.2(2.6) |
| CCU | 2.1(0.4) | 3.0(0.6) | 2.6(0.4) | 2.8(0.5) | 2.6(0.7) | 1.7(0.4) | 1.7(0.4) | 1.9(0.6) | 2.3(0.4) |

### 3.3 Voxel-wise comparison against multicenter data

We also validated our method with multi-center data, which includes data from Charité – Universitätsmedizin Berlin (CUB), University of California, Los Angeles (UCLA), and publicly available data from Johns Hopkins University (JHU).

DSC were computed between manually corrected segmentation using the SAMIRIX toolbox and the proposed method’s outcome for 9 classes for CUB and UCLA. The corresponding values are reported in Table 5, where vitreous is the region above ILM and b-BM is the region below BM, as shown in Figure 5. As it can be seen from Table 5, the first and the last classes (vitreous, b-BM) are with the highest DSC values (close to 1) as the difference between numbers of background and foreground pixels are small. Moreover, OPL has the lowest value of DSC since this is one the thinnest layers and the difference between numbers of background and foreground pixels can be large. Compared to the state-of-the-art methods, our method produces better overall DSC scores (CUB-0.95, UCLA-0.94, and JHU-0.92) except Borkovkina et al. (2020) and Ngo et al. (2019), where validations are performed using HC data only.

**Table 5:**
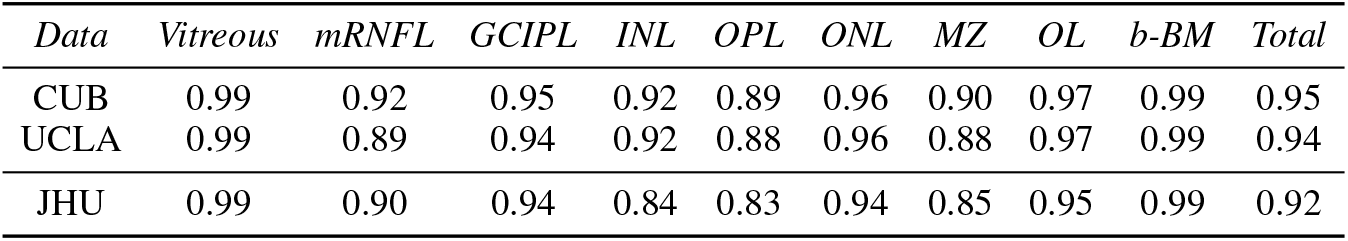
DSC between CCU-INSEG’s outcome and the manually segmented data.

| *Data* | *Vitreous* | *mRNFL* | *GCIPL* | *INL* | *OPL* | *ONL* | *MZ* | *OL* | *b-BM* | *Total* |
| --- | --- | --- | --- | --- | --- | --- | --- | --- | --- | --- |
| CUB | 0.99 | 0.92 | 0.95 | 0.92 | 0.89 | 0.96 | 0.90 | 0.97 | 0.99 | 0.95 |
| UCLA | 0.99 | 0.89 | 0.94 | 0.92 | 0.88 | 0.96 | 0.88 | 0.97 | 0.99 | 0.94 |
| JHU | 0.99 | 0.90 | 0.94 | 0.84 | 0.83 | 0.94 | 0.85 | 0.95 | 0.99 | 0.92 |

**Figure 5:**
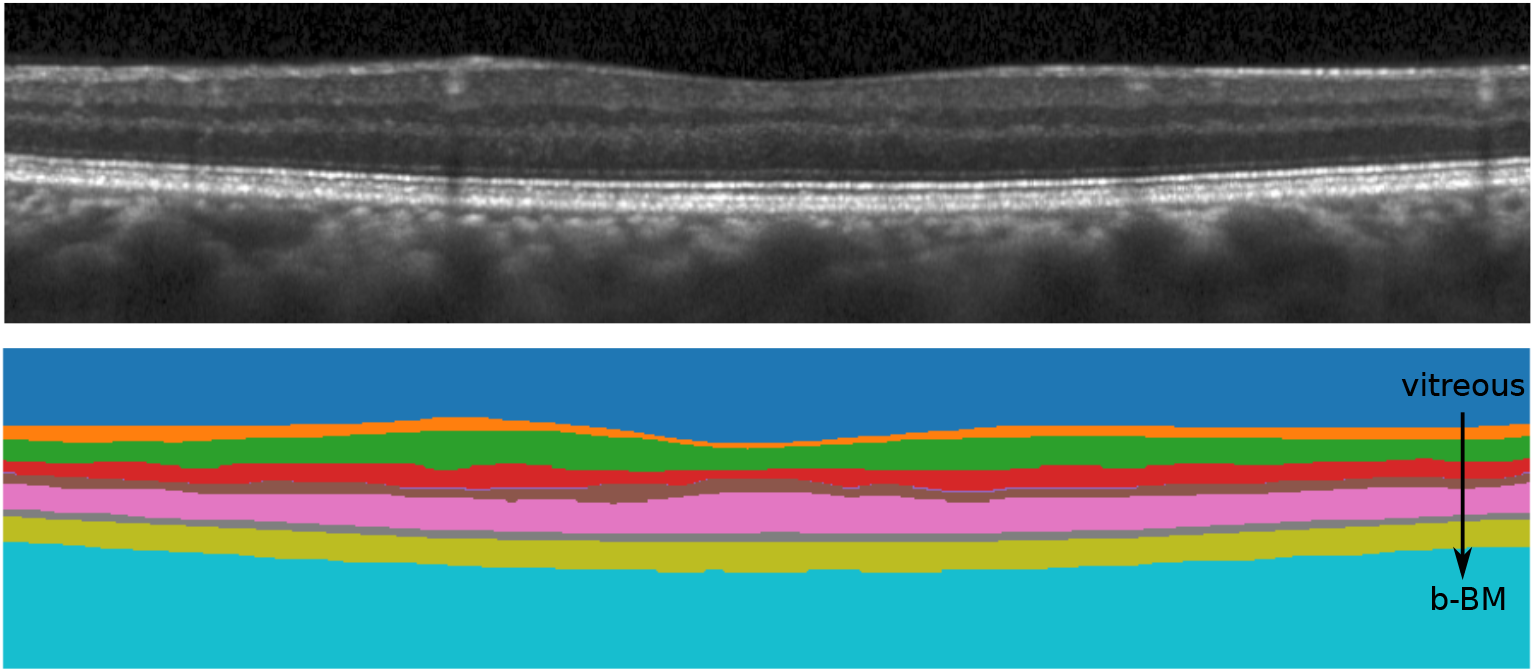
All nine different classes segmented by the proposed method.

Table 6 shows layer-based MAE computed between CCU-INSEG’s outcome and the manually corrected gold standard segmentation for the CUB and UCLA data sets and the manually corrected segmentation by the publishers for the JHU data set. As it can be seen, the proposed method produces a high fidelity segmentation with regard to the gold standard segmentation. Errors for different layers are in sub-pixels, except for the INL-OPL boundary in the JHU data. Furthermore, an experienced layers segmentation expert examined all segmented B-scans and rejected **1.5%, 4.7%**, and **2.8%** B-scans from CUB, UCLA, and JHU data respectively.

**Table 6:** MAE (standard deviation) in *μ*m between CCU-INSEG’s outcome and the manually corrected segmentation for each boundary.

| *Data* | *ILM* | *RNFL-GCL* | *IPL-INL* | *INL-OPL* | *OPL-ONL* | *ELM* | *EZ-OS* | *BM* | *Total* |
| --- | --- | --- | --- | --- | --- | --- | --- | --- | --- |
| CUB | 2.1(0.4) | 3.1(0.6) | 2.5(0.4) | 2.9(0.4) | 2.6(0.7) | 1.8(0.4) | 1.7(0.4) | 1.8(0.4) | 2.3(0.4) |
| UCLA | 2.2(0.2) | 4.2(1.0) | 2.7(0.4) | 3.2(0.5) | 2.9(0.4) | 2.0(0.3) | 2.0(0.4) | 2.0(0.5) | 2.6(0.3) |
| JHU | 3.2(0.4) | 4.0(0.7) | 3.9(0.6) | 5.3(1.0) | 3.8(1.0) | 2.6(0.4) | 2.6(0.4) | 4.0(0.9) | 3.7(0.3) |

We would like to emphasize an important point here that our method is trained only on the data from one center (CUB). The CCU network was not trained on the data either from UCLA or from JHU. However, the performance of our method is quite similar for all these data in terms of MAE (sub-pixel level). Figure 6 shows the effectiveness of the proposed method against different challenges from these three different data sets. Figure 6 (top) is a noisy scan (CUB data), Figure 6 (middle) is a scan with microcysts from JHU data, and UCLA data has a quite thin RNFL on one side of the macula. As it can be seen from Table 6, MAE for BM and INL-OPL are a bit on higher side (close to 1 pixel) in JHU data, where manual segmentation was not performed by the same expert as CUB and UCLA, and it is quite possible that the segmentation from different experts are not exactly the same (Oberwahrenbrock et al. (2018)).

**Figure 6:**
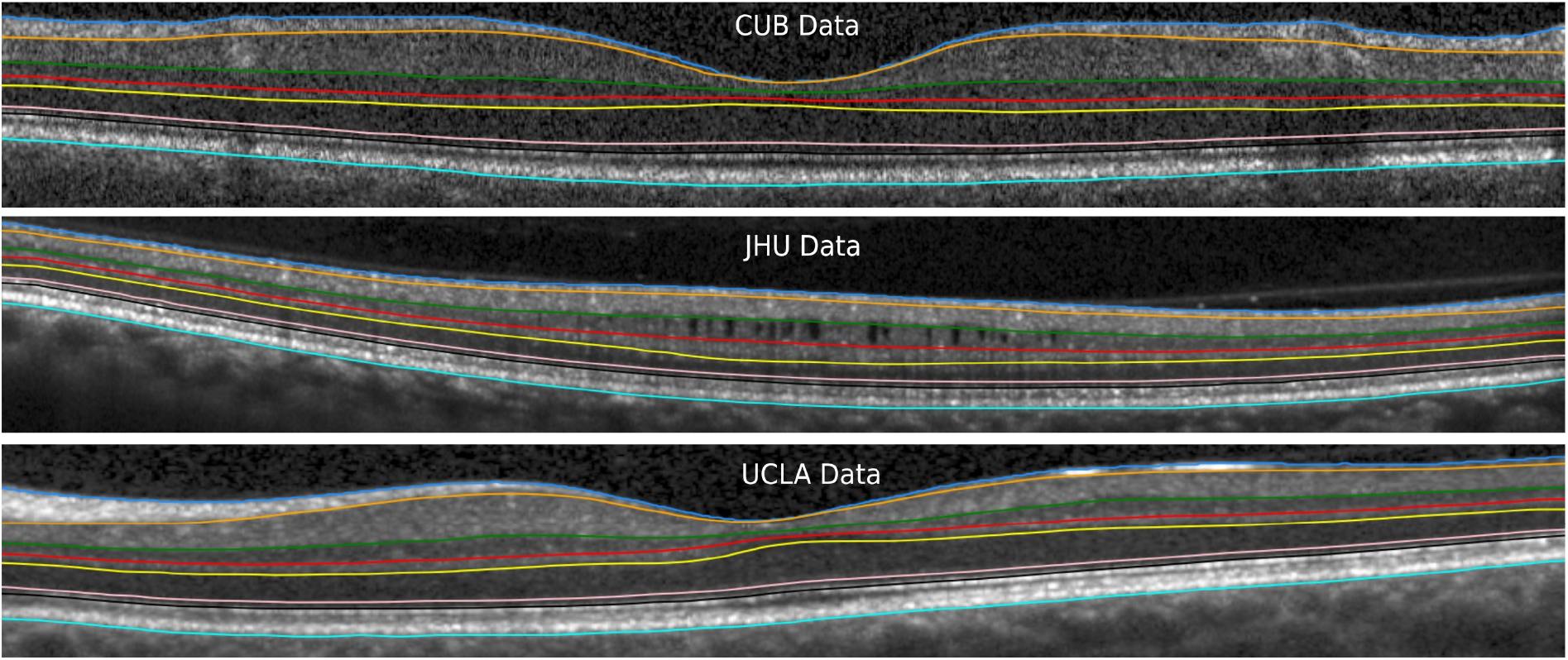
The visualization of the segmentation lines computed by the proposed method for three different data sets.

### 3.4 Reliability and fidelity of derived thickness and volume parameters

In this section, we compute the standard OCT parameters based on the segmented layers using the proposed method. Here, we are focusing on five standard thickness parameters within a 5mm circular region from the foveal center. The parameters are described in Figure 2, and additionally we provide data for the ganglion cell complex (GCC, tissue between ILM and IPL-INL boundary), and total macular thickness (TM, between ILM and BM).

The test-retest reliability of the proposed method is performed on a data set, which has three repeated measurements of 30 healthy eyes. Intra-class correlation coefficient (ICC) for the standard parameters varied from 0.93 (mRNFL) to 0.99 (GCIPL, GCC, and total macula) for both thicknesses and volumes as shown in Table 7. As shown in Table 7, mRNFL has the lowest ICC because mRNFL is one of the thinnest layers in the retina. Therefore, the measurement of this layer can be easily contaminated by noise components, which lead to the lower value of ICC.

**Table 7:** The above table shows reliability of the standard parameters (top three row) and fidelity of the computed parameters (MAE in *μ*m) from CCU-INSEG’s segmentation with respect to the manual segmentation (the last three rows). Abbreviations (thick. - thickness, vol. - volume, ICC - intra-class correlation coefficient, LCI - lower confidence interval, UCI - upper confidence interval, mRNFL - Macular retinal nerve fiber layer, GCIPL - Macular ganglion cell and inner plexiform layer, INL - Inner nuclear layer, GCC - Ganglion cell complex, TM - Total macula.)

| *Param* | *mRNFL*<br>*thick.* | *GCIPL*<br>*thick.* | *INL*<br>*thick.* | *GCC*<br>*thick.* | *TM*<br>*thick.* | *mRNFL*<br>*vol.* | *GCIPL*<br>*vol.* | *INL*<br>*vol.* | *GCC*<br>*vol.* | *TM*<br>*vol.* |
| --- | --- | --- | --- | --- | --- | --- | --- | --- | --- | --- |
| ICC | 0.93 | 0.99 | 0.96 | 0.99 | 0.99 | 0.93 | 0.99 | 0.96 | 0.99 | 0.99 |
| LCI | 0.88 | 0.98 | 0.93 | 0.97 | 0.98 | 0.88 | 0.98 | 0.93 | 0.97 | 0.98 |
| UCI | 0.96 | 0.99 | 0.98 | 0.99 | 0.99 | 0.96 | 0.99 | 0.98 | 0.99 | 0.99 |
| CUB | 1.2 | 0.9 | 0.8 | 0.5 | 0.7 | 0.024 | 0.018 | 0.016 | 0.010 | 0.014 |
| UCLA | 2.0 | 1.6 | 0.8 | 0.5 | 0.6 | 0.039 | 0.032 | 0.016 | 0.009 | 0.013 |
| JHU | 0.9 | 1.8 | 6.9 | 1.4 | 2.4 | 0.018 | 0.035 | 0.140 | 0.028 | 0.044 |

To check the fidelity of our method, we use the same multi-center data sets with the gold standard segmentation. Furthermore, the standard parameters are computed on both segmentation (the gold standard and the outcome of the proposed method). Table 7 (the last three rows) shows MAEs computed between parameters values based on gold standard segmentation and our outcome. For CUB and UCLA data, the errors are limited up to 2 *μ*m for thickness parameters. For JHU data, INL related parameters are not quite similar. Perhaps, the higher error is because of the different manual corrections at different centers.

### 3.5 Comparison against state-of-the-art methods

For comparison against state-of-the-art methods we randomly selected 20 macula volume scans of 20 eyes from patients with NMOSD from a recent publication (Motamedi et al. (2019)). Eyes from NMOSD patients regularly show severe neuroaxonal retinal damage, resulting in problematic image quality and/or larger neuroaxonal changes. While this does not reflect performance in a random data set, it does test segmentation accuracy in difficult data. For comparison, we chose AURA (a graph-based segmentation method) (Lang et al. (2013)), and deep learning-based RelayNet (Roy et al. (2017)) for their availability.

First, we presented segmentation results to an independent experienced grader (EMK) in a blinded fashion. The expert rejected **41.4% (317 from 958 images)** B-scans segmented by AURA, but only **3.5 % (34 from 958 images)** B-scans from our proposed method (Fig. 7).

**Figure 7:**
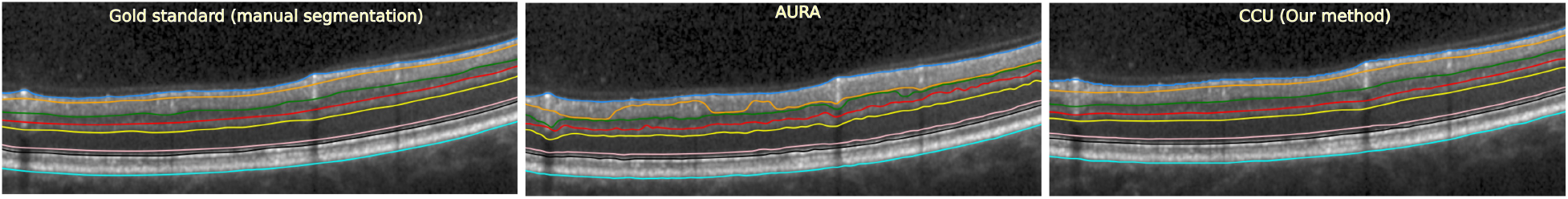
Example segmentations from AURA (Lang et al. (2013)) and CCU (Our method).

Unfortunately, a similar comparison is not possible with RelayNet (Roy et al. (2017)) as this method provides only class pixels (Note: we are using the PyTorch implementation). Therefore, we performed the comparison with RelayNet based on DSC using 45 manually delineated B-scan images and performed the segmentation using AURA, RelayNet, and CCU (Fig. 7).

Table 8 shows that our methods outperforms the state-of-the-art methods in this problematic data set. AURA performs weak for mRNFL and GCIPL, which as been reported before (He et al. (2019)). RelayNet produced several false positive and false negative pixels in the prediction leading to overall low DSC values.

**Table 8:** DSC between CCU, AURA and RelayNet. ⋆ RelayNet segments ONL + MZ in a single class, therefore MZ column is empty for RelayNet and ONL column shows DSC for both ONL and MZ. RelayNet segments OL in two different classes. For this comparison, we combined both classes in OL and computed DSC with respect to manual segmentation.

| *Method* | *Vitreous* | *mRNFL* | *GCIPL* | *INL* | *OPL* | *ONL* | *MZ* | *OL* | *b-BM* | *Total* |
| --- | --- | --- | --- | --- | --- | --- | --- | --- | --- | --- |
| AURA (Lang et al. (2013)) | 0.99 | **0.57** | **0.62** | 0.86 | 0.85 | 0.96 | 0.92 | 0.98 | 0.99 | **0.86** |
| RelayNet (Roy et al. (2017)) | 0.87 | 0.70 | 0.73 | 0.64 | 0.86 | 0.77* | - | 0.90 | 0.92 | **0.80** |
| CCU | 0.99 | **0.88** | **0.93** | 0.88 | 0.84 | 0.96 | 0.89 | 0.98 | 0.99 | **0.93** |

## 4 Discussion

In this paper, we propose a method for automatic intraretinal layer segmentation of macular OCT images (CCU-INSEG). The proposed method is a two-stage segmentation algorithm, where the retinal region is separated in the first stage and intra-retinal layer segmentation is performed in the second stage. The focus of this work was to develop an efficient and high fidelity segmentation algorithm, which can be easily deployed on a device with limited resources (in terms of memory and computational power). Therefore, we performed distillation on the original U-Net architecture and propose a compressed version of U-Net with comparable accuracy. We cascaded two different compressed versions of U-Net in the proposed method and these networks are 392x and 26x smaller than the original U-net, respectively (Table 2).

Our study demonstrates that good segmentation accuracy can be achieved using less sophisticated networks, but meaningful domain-specific processing and training data selection. We selected a heterogeneous training data set, which included healthy controls and different autoimmune inflammatory and neurodegenerative diseases of the central nervous system. Thorough quality control was performed on each image, and we selected images representing a range of real-world quality including low quality images. Nonetheless, like previous methods, our method is still hyperspecialized towards retinas without larger pathological changes. As a consequence, segmentation of structural pathologies from eye disorders such as AMD will most likely fail. Improving representativeness of training data including class expansions are certain ways to further improve this and any other method towards clinical application.

Staurenghi et al. (2014) proposed a lexicon for uniform naming of anatomical structures based on a panel consensus. This consensus was adapted as part of the APOSTEL recommendations for OCT studies (Cruz-Herranz et al. (2016)), and we here utilize this lexicon for structural labels. De Fauw et al. (2018) provided a consensus for naming pathological structures from multiple common eye disorders in their landmark artificial intelligence study. However, both nomenclatures are not widely adopted, and there is an unmet need for a widely supported lexicon of anatomical and pathological findings, which would serve as a guide for OCT classification in artificial intelligence applications, and would also improve the sustainability of published data sets and methods.

Voxel-wise performance evaluation of our proposed method is done on multi-center data, which included data from three different centers (CUB, UCLA, and JHU). We have shown in Table 6 that our method produces almost equally good segmentation (sub-pixel level MAE) with reference segmented data from these centers. Importantly, one data set was from patients with glaucoma, and this disease was not part of the training data set. Nonetheless, performance was comparable, confirming that our method can extend beyond its training domain. On the other hand, while generally good, the noise was higher than expected on validation against JHU data, mainly for INL-OPL boundary (above a pixel) and BM (close to a pixel). We hypothesize that this was caused by different graders manually correcting each data set. Previously, we have shown that manual grader segmentation can systematically differ within and between centers, which is one of the core motivations for a fully automatic, reproducible segmentation method (Oberwahrenbrock et al. (2018)).

Evaluation standards for retinal segmentation methods are lacking, which is shown by different validation approaches used by the methods summarized above in state of the art. Tian et al. (2016) have proposed methods for how to perform evaluation, but there is a need for unbiased performance evaluations of automated segmentation algorithms using e.g. real-world representative data sets with an independent consensus ground truth.

While outcomes of the proposed method are encouraging for neurology and ophthalmology clinical and research applications, ours and other proposed methods are still limited in their generalizability. While many methods may work in specific limited scenarios, e.g. in a research setting, wide clinical application requires extension of supported devices, retinal features, handling of longitudinal data and more.

## Data Availability

All data produced in the present study are available upon reasonable request to the authors. Due to EU GDPR data privacy regulations, OCT source data cannot be shared or made publicly available.

